# Weight trajectories after last Tirzepatide or Semaglutide prescription across a federated health network

**DOI:** 10.64898/2026.01.26.26344839

**Authors:** Karthik Murugadoss, Gowtham Varma, AJ Venkatakrishnan, Michael C. Gibson, Venky Soundararajan

## Abstract

GLP-1 receptor agonist (GLP-1RA) discontinuation has been associated with weight regain. However, the real-world association between discontinuation of GLP-1RA prescriptions and weight change has not been explored. We assessed weight trajectories of 4,182 patients in the six months following their last GLP-1RA prescription. Approximately two-thirds of patients showed stable weight or continued weight loss during this period post the last known GLP-1RA prescription. In a representative subset of patients with clinician-documented discontinuation near the last prescription (N=300), a similar distribution of weight regain in a minority of patients was observed vs no regain in the majority of patients during the six-month post-GLP-1RA prescription period. To mirror clinical trial-style discontinuation definitions, we also evaluated cohorts with no subsequent GLP-1RA prescription for 1 year after the last prescription (semaglutide N=1,755; tirzepatide N=1,312), observing weight regain in a minority of patients (39.3% semaglutide; 26.6% tirzepatide) and no weight regain in the majority (60.7% semaglutide; 73.4% tirzepatide) in the year following the last known GLP-1RA prescription. Exercise counseling was documented more frequently among patients with durable weight loss post-last GLP1 prescription compared with those with weight regain (26.2% vs. 14.7%; p=0.04). Further studies are warranted to infer the mechanisms underlying these real-world patterns.

---

GLP-1 receptor agonists (GLP-1RAs) such as semaglutide and tirzepatide have transformed obesity care and weight management, delivering unprecedented levels of weight loss in real-world practice.^1^ Several studies have demonstrated reversal of weight loss and other cardiometabolic benefits after patients discontinue GLP-1RAs.^2–4^ However, it is not clear whether there are subsets of patients who are more likely to experience continued benefit after their therapeutic exposure. This question has recently been further complicated by the trend for patients to obtain GLP-1RA prescriptions from other sources such as online providers or medical spas.^5^ Thus, at the intersection of the patient and clinician experience, it is important to better understand patient trajectories following the last GLP-1RA prescription by a primary care or specialist physician.

Using large-scale longitudinal real-world data on the nSights platform, we analyzed weight trajectories before and after the last GLP-1RA prescription to directly address this question (**Figure 1A**). Specifically, among patients who had at least three GLP-1RA prescriptions, we assessed the average percent change in weight in the year leading up to their last prescription (defined as month 0) and in the six months following their last prescription. Separate analyses were performed for semaglutide and tirzepatide.

**Figure 1.**
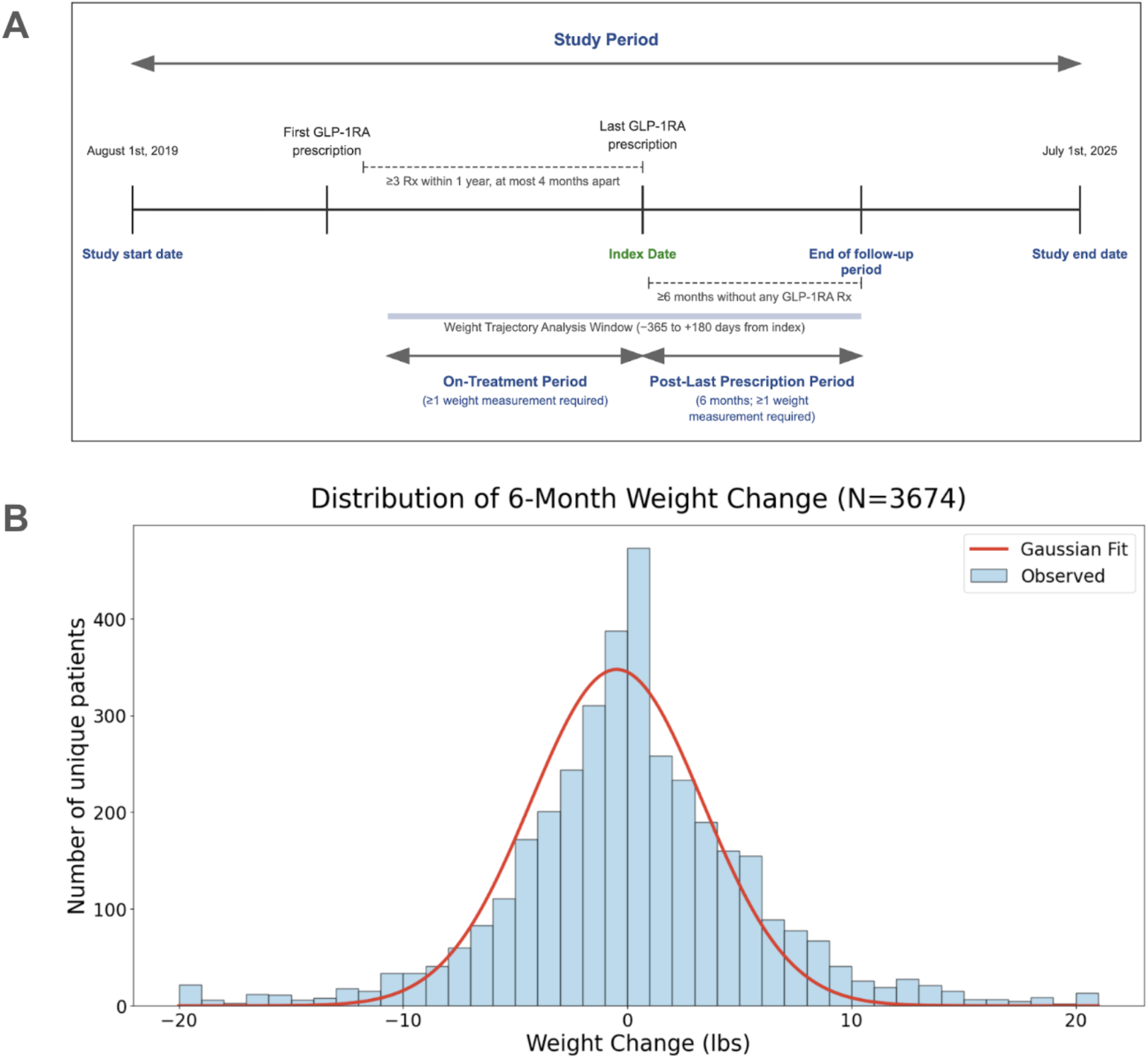
(A) Study schematic. Weight trajectories of patients with at least three prescriptions of semaglutide or tirzepatide were assessed for six months following the last prescription. (B) Histogram depicting the distribution of weight change during the follow-up period. For each individual patient, the weight measurements that were chronologically closest to the index date (date of late prescription) and six months after the index date were used to calculate the patient-level weight change.

There were 2,567 patients in the semaglutide cohort and 1,615 patients in the tirzepatide cohort. Demographic and baseline clinical characteristics are summarized in **Table 1**. Briefly, the semaglutide cohort was about 71.2% female and 77.3% white with a mean age of 55 years (SD: 13 years); the tirzepatide cohort was about 70.2% female, 85.3% white, and had a mean age of 55 years (SD: 12 years). At the time of the last prescription, the prevalence of type 2 diabetes and obesity, respectively, were 60.0% and 66.0% in the semaglutide cohort, and 77.0% and 82.5% in the tirzepatide cohort. Average weight in the semaglutide and tirzepatide cohorts were 93.3 kg and 93.4 kg, respectively, and average BMI were 32.0 kg/m^2^ and 31.4 kg/m^2^, respectively. At least one prescription for a recommended maintenance dose (see **Methods**) during the year prior to the last prescription was present for 90.6% of patients taking semaglutide and 95.3% of patients taking tirzepatide.

**Table 1.** Demographics and baseline clinical characteristics for the semaglutide and tirzepatide cohorts.

|  | **Semaglutide Cohort**<br>**(N = 2,567)** | **Tirzepatide**<br>**(N = 1,615)** |
| --- | --- | --- |
| **Age** |  |  |
| Age at last prescription (Mean, Std) | 55.3 (12.8) | 55.1 (11.8) |
| **Sex** |  |  |
| Female (N, %) | 1,828 (71.2%) | 1,134 (70.2%) |
| Male (N, %) | 739 (28.8%) | 481 (29.8%) |
| **Race** |  |  |
| White (N, %) | 1,984 (77.3%) | 1,378 (85.3%) |
| African American / Black (N, %) | 421 (16.4%) | 131 (8.1%) |
| Other (N, %) | 162 (6.3%) | 106 (6.6%) |
| **Clinical Characteristics** |  |  |
| Type 2 Diabetes Mellitus (N, %) | 1,540 (60.0%) | 1,066 (66.0%) |
| Obesity (N, %) | 1,977 (77.0%) | 1,332 (82.5%) |
| Weight, kg (Mean, SD) | 93.3 (22.4) | 93.4 (23.5) |
| BMI, kg/m<sup>2</sup> (Mean, SD) | 32.0 (5.5) | 31.4 (6.2) |

The distribution of average weight change after the last GLP-1RA prescription is approximately a normal distribution with a mean value of 0.29% (median value of 0), indicating that most patients did not experience significant weight gain following their last GLP-1RA prescription (**Figure 1B**). Patients were grouped retrospectively based on their post-last prescription trajectories into two clinically intuitive categories: (1) patients with at least 2% increase in weight in the six months following their last GLP-1RA prescription (“weight regain”); (2) patients without at least 2% increase in weight following their last GLP-1RA prescription (“non-regain”). In both the tirzepatide- and semaglutide-treated cohorts, more than two-thirds of patients fell into the latter group without weight regain after the last prescription (**Figure 2**). Of note, the weight loss trends in the year leading up to the last prescription were quite similar between the weight regain and non-regain groups for both semaglutide (∼7% weight loss) and tirzepatide (∼11% weight loss). At the time of the last prescription, the weight and BMI distributions in the two groups were also similar for both medications (**Figure 3**). Among patients who regained weight after their last GLP-1RA prescription (33.0% for semaglutide, 27.6% for tirzepatide), the average weight regain reached 4% within four months of the last prescription (**Figure 2**). In a subset of 139 patients with a GLP-1RA prescription that was explicitly discontinued, 34% of semaglutide patients and 30% of tirzepatide patients experienced weight regain in the next six months.

**Figure 2.**
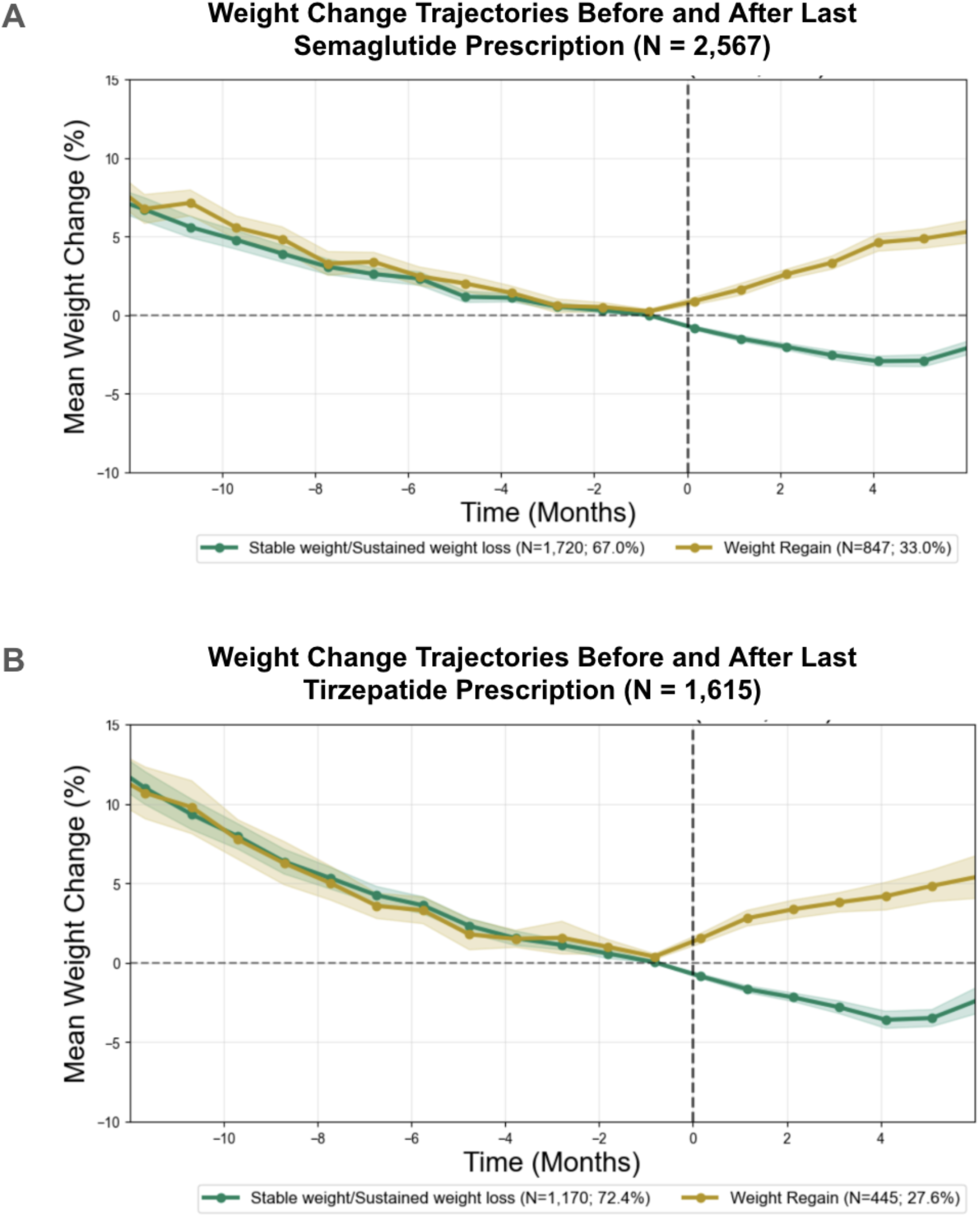
Weight loss trajectories of patients leading up to and after the last prescription of semaglutide (A) or tirzepatide (B), divided into subsets of patients who demonstrated weight regain (orange) versus stable weight or continued weight loss (green).

**Figure 3.**
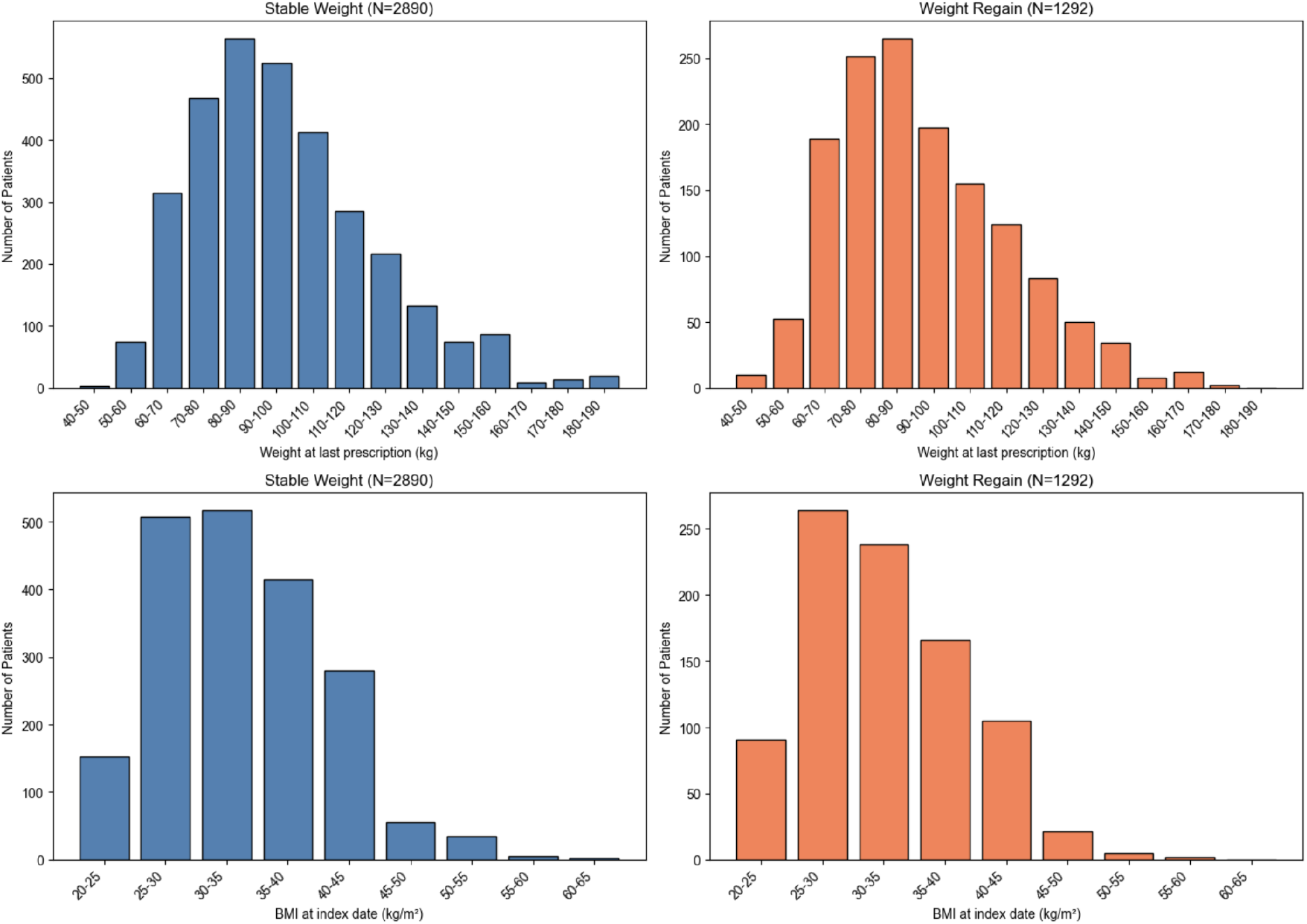
Weight (top row) and BMI distributions (bottom row) for the cohorts of patients with stable weight following the last GLP-1RA prescription (left column) or weight regain following the last prescription (right column). The weight and BMI closest to the date of the last prescription were selected for each patient.

Consistent with a clinical trial-style discontinuation proxy, restricting to patients with no subsequent GLP-1RA prescription for 1 year after the last prescription (semaglutide N=1,755; tirzepatide N=1,312) similarly showed weight regain in a minority of patients (39.3% semaglutide; 26.6% tirzepatide) and no weight regain in the majority (60.7% semaglutide; 73.4% tirzepatide) in the year following the last known prescription.

We next evaluated whether lifestyle factors, specifically diet and physical activity, might be associated with sustained weight maintenance. At a representative site, we applied large language models (LLMs) to analyze the frequency of lifestyle counseling documented in longitudinal electronic health record (EHR) notes (see **Methods**) among patients who did not experience weight regain (N = 226) versus those who did (N = 74). Patients without weight regain were more likely to have exercise counseling mentions than those with regain (26.2% vs. 14.7%; p=0.04; **Figure 4**). In contrast, rates of diet counseling after the last prescription were similar between these groups (22.3% vs. 23.5%; p=0.83; **Figure 4**).

**Figure 4.**
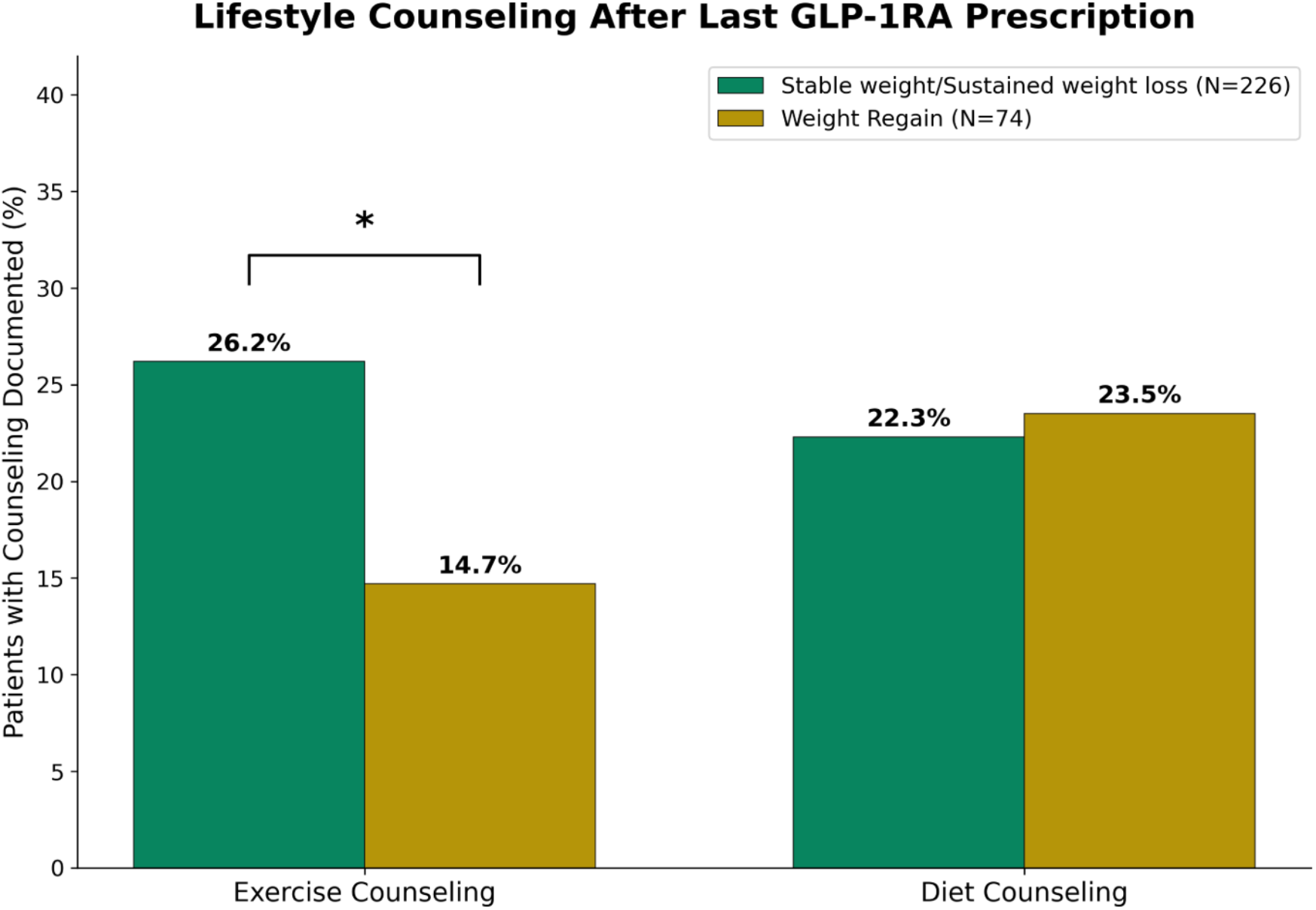
Comparison of rates of documented lifestyle counseling in the period following the last GLP-1RA prescription, including exercise counseling and diet counseling. All patients from a single health system within the federated network (N = 300) were included in this analysis. Statistical significance: *, p < 0.05.

One limitation of the current study design is that the analysis is anchored on the date of each patient’s last prescription, which does not necessarily correspond to medication discontinuation. To better understand the relationship between these events, we deployed a large language model across clinical notes to determine how frequently there was supporting clinician documentation of treatment cessation within ±90 days of the last prescription (see **Methods**). There was such documentation for 39.5% and 44% of the weight regain and non-regain groups for the semaglutide cohort, respectively, and for 51.6% and 31.8% of these groups for the tirzepatide cohort (**Table 2**). It was rare to find notes indicating that patients subsequently obtained the same or another GLP-1RA from another source (**Table 2**), although this likely underestimates the true prevalence of such behavior due to incomplete reporting and documentation.

**Table 2.**
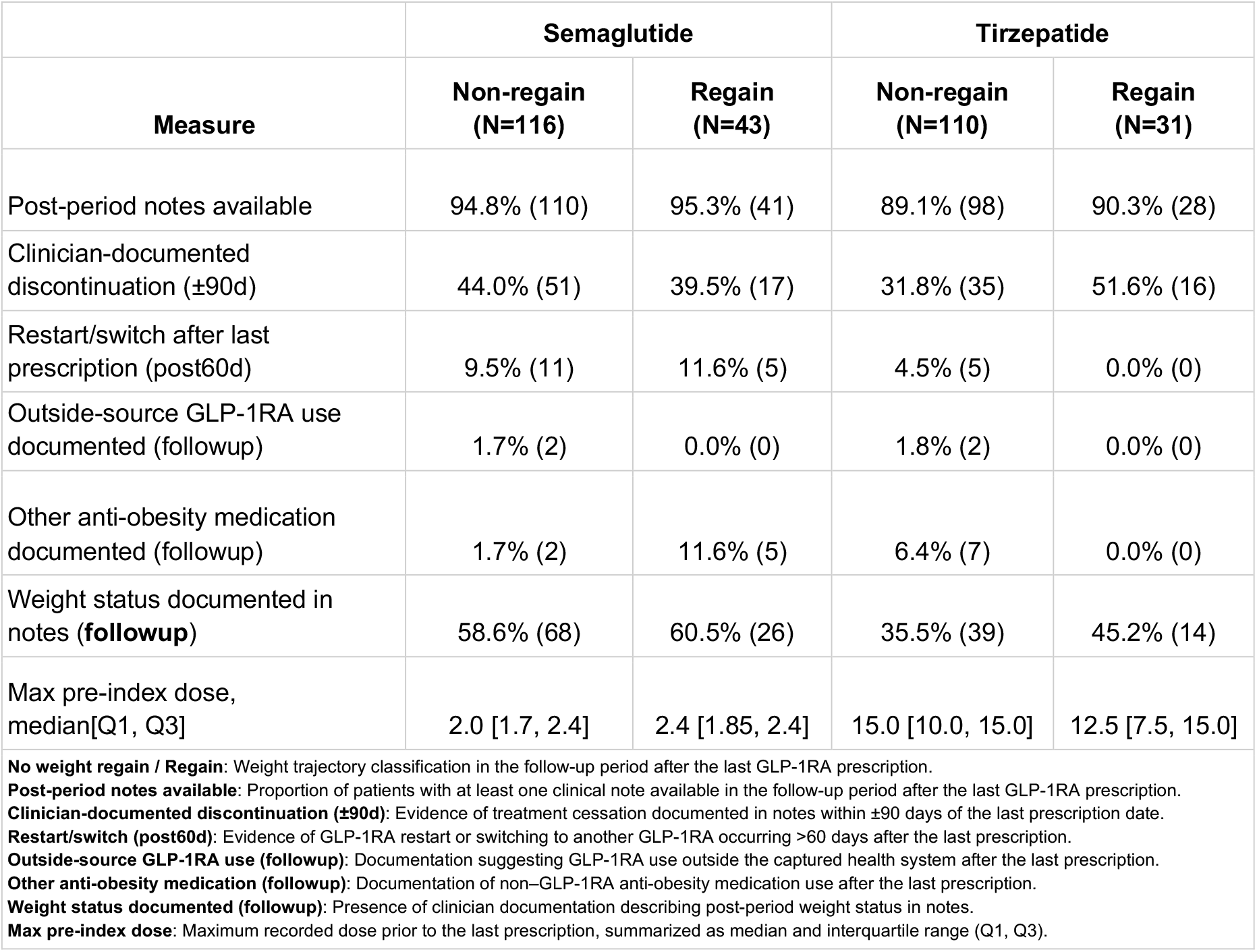
Summary of LLM-extracted content from clinical notes in the analyzed cohorts, split by both medication (semaglutide versus tirzepatide) and weight trajectory in the six months following the last prescription (weight regain versus non-regain). All patients from a single health system within the federated network (N = 300) were included in this analysis.

Overall, this study highlights that while patients with an active GLP-1RA prescription tend to lose significant weight, weight regain is not universally observed after discontinuation of the prescription. There are likely multiple factors contributing to the observation that most patients had stable weight or continued weight loss after the last prescription in this study. For example, as described above, exercise counseling was documented more frequently in the non-regain cohorts. This suggests that lifestyle factors such as exercise may play an important role in modulating weight loss durability following the last GLP-1RA prescription and motivates randomized clinical trials to directly test this hypothesis. Further, some patients may have continued to have treatment exposure following the date of the last prescription, whether related to this prescription itself or medication access from another source such as a medical spa, telehealth provider, and/or compounding pharmacy. Finally, there could be sustained pharmacological benefit in some patients that persists even months after semaglutide or tirzepatide has been discontinued. In any case, the findings of this study can help both patients and clinicians better understand what happens in the real world after a primary care or specialist physician stops prescribing a GLP-1RA.

For multiple reasons, the data presented here is not contradictory to prior randomized controlled trials (RCTs) that have demonstrated weight regain after discontinuation of GLP-1RAs.^2,4,6,7^ First, this analysis focuses on weight trajectories that follow the last GLP-1RA prescription by a primary care or specialist physician, a unique “real-world index event” that has not been explored in previous studies. Further, RCTs have measured weight rebound under controlled withdrawal settings (e.g., withdrawal at a specified timepoint after therapy initiation), whereas real-world data reflects a combination of pharmacology, adaptive human behavior, selective discontinuation, flexible re-initiation patterns, and more. Indeed, real-world treatment cessation is heterogenous and often partial, staggered, or followed by re-initiation, dose tapering, lifestyle changes, or adjunctive therapies, all of which could blunt or magnify metabolic rebound physiology.^8,9^ Patients who discontinue therapy and do not restart within the following six months, as is the case for the cohorts analyzed here, are by definition a selected subset who likely have unique characteristics relating to treatment exposures and behavioral adaptations. Consistent with this interpretation, applying a more stringent clinical trial-style discontinuation proxy of no subsequent GLP-1RA prescription for 1 year after the last prescription yielded a similar qualitative pattern, with weight regain occurring in a minority of patients and weight stability or continued loss in the majority. The implication of this study is not that the risk of weight rebound is negligible but rather that durable response after prescription discontinuation may be achievable with an appropriate framework in place. This argues for emphasizing a longitudinal disease management paradigm for obesity pharmacotherapy, where agentic care pathways can help clinicians better identify who can safely discontinue, who requires continued maintenance therapy, and when sustained intervention is indicated to preserve metabolic benefits.

This study has several limitations. It is a retrospective analysis of real-world data and is thus prone to multiple sources of confounding. Second, the follow-up interval is limited to 6 months, whereas prior studies have explored longer durations following treatment cessation.^3,4,6,10^ The shorter-interval follow-up could lead to underestimation of patients who regain weight after the last prescription. To partially address this limitation, we conducted a secondary analysis restricted to patients with no subsequent GLP-1RA prescription for 1 year after the last prescription, enabling assessment of regain versus non-regain over a longer post-prescription interval. Studies with longer-term follow-up will become more feasible in the coming months. Third, there is certainly variability in treatment adherence, even leading up to the last prescription. To enrich for patients who were more likely to have had true therapy exposure, we required patients to have at least three GLP-1RA prescriptions at most four months apart during a one-year period in the participating health systems for inclusion in the study. However, it is likely that some of these patients missed doses or stopped taking the medication before the last documented prescription. Finally, this study did not account for other medical comorbidities and social determinants of health including insurance status, which are known to impact access to therapies including prescription fill rates of GLP-1RAs specifically.^11^ Future analyses are warranted to explore how these factors impact weight gain in the post-GLP-1RA prescription interval.

In conclusion, weight loss after GLP-1RA therapy does not uniformly reverse after prescriptions are discontinued. In real-world data, a majority of patients maintain or continue their weight loss, while a smaller but important subset experiences weight regain. These findings illustrate the importance of a nuanced view of long-term weight regulation in the era of GLP-1 therapies and motivate future multimodal monitoring of patients after prescription discontinuation and/or therapy cessation which integrate biology, behavior, comorbidities, and drug dosing information. Such investigations could unlock features that can help to predict patients who are likely to show sustained benefit versus rebound of weight and other cardiometabolic parameters.

## Methods

### Study Design and Patient Population

This was a retrospective observational study to assess patterns of weight change (“weight trajectories”) following the last GLP-1RA prescription by a provider within a federated system of multiple health networks. Patients were eligible for inclusion in the study if they had (1) at least three prescriptions for semaglutide or tirzepatide at most 4 months apart within a one-year period between 08/01/2019 to 07/01/2025, (2) subsequently had at least six months without a prescription for any GLP-1RA medication, and (3) had at least one weight measurement during their on-therapy time and at least one weight measurement within six months after the date of their last GLP-1RA prescription. Patients were excluded if they ever had a prescription for another GLP-1RA therapy, in order to limit the potential of confounding due to treatment-switching effects. For each patient, the index date was set as the date of their last GLP-1RA prescription, specifically the date of the GLP-1RA prescription that was followed by at least six months without another prescription for the same medication. In a secondary, more stringent analysis intended to mirror clinical trial-style discontinuation, we further restricted to patients with no subsequent GLP-1RA prescription for 1 year after the index date and evaluated weight regain versus non-regain over that 1-year post-index interval.

### Analysis of Weight Trajectories

Weight measurements were analyzed from 12 months before the last prescription through 6 months after the last prescription (−365 to +180 days from the index date). For each measurement, percentage weight change was calculated as the difference from baseline weight divided by baseline weight, multiplied by 100. Measurements were then grouped into 30-day bins relative to the index date. To characterize cohort-level weight trajectories while accounting for variable measurement frequency across patients, a two-stage aggregation approach was employed. First, patient-level median weight change was calculated within each time bin to generate at most a single representative value per patient per bin. Cohort-level statistics, including mean and standard deviation, were then derived from these patient-level values. Ninety-five percent confidence intervals were computed as mean ± 1.96 × standard error of the mean, where standard error of the mean was defined as the standard deviation divided by the square root of the number of patients contributing to each bin.

### Definition of Recommended Maintenance Doses

We evaluated maintenance dose attainment within the 12 months preceding (and including) the date of last prescription. Maintenance doses were defined as the minimum labeled therapeutic dose for each formulation: 0.5 mg for Ozempic, 1.7 mg for Wegovy, 7 mg for Rybelsus, and 5 mg for Mounjaro and Zepbound. A patient was classified as having attained maintenance dosing if at least one prescription during this window met or exceeded the threshold for their corresponding medication. The percentage of patients reaching maintenance dose was calculated separately for semaglutide and tirzepatide, with the denominator comprising all patients with sufficient follow-up data for trajectory classification.

### Content Extraction from Clinical Notes

We applied a large language model (LLM) with schema-constrained structured decoding to extract GLP-1 treatment context from longitudinal clinical notes for all study patients at one of the academic medical centers within the federated network. For each patient, we selected notes within 12 months before and after the index date (last prescription) that contained a generic or brand mention of semaglutide or tirzepatide. Extraction was performed using GPT-OSS 20B, deployed via vLLM (version 0.13.0) in Python (version 3.13), with an average input token length of 3.5k tokens per note. Inference was conducted with temperature 0.1 and top_k=1 using a fixed random seed to ensure reproducibility. The LLM produced note-level JSON outputs capturing (1) clinical-documented GLP-1 status changes (e.g., held, stopped, restarted, switched) and dosing information, (2) lifestyle counseling (diet and exercise), (3) use of other anti-obesity medications, and (4) optional context such as access barriers or outside sourcing when explicitly stated. All extracted events required short verbatim evidence excerpts from the note to support provenance, and note-level extractions were subsequently aggregated into patient-level indicators relative to the index date for downstream cohort summaries and comparisons across outcome groups.

### Statistical Analysis

Differences in the frequency of lifestyle counseling documentation between patients with and without weight regain were evaluated using chi-square tests for categorical variables. Two-sided p-values less than 0.05 were considered statistically significant. Analyses were performed separately for diet counseling and exercise counseling mentions extracted from clinical notes documented after the last GLP-1RA prescription. All statistical analyses were conducted using Python (version 3.13) with the SciPy library (version 1.15.1).

### Data source & selection

This study analyzed de-identified EHR data from a network of tertiary clinical centers tied to academic medical centers in the United States through the nference nSights Analytics Platform. nference, in collaboration with academic medical center (AMC) data partners, provided the de-identified data for this study. nference has established a secure data environment, hosted by and within each of the AMCs, that contain the AMC’s de-identified patient data. The provisioning of and access to this data are governed by an expert determination that satisfies the HIPAA Privacy Rule requirements for the de-identification of protected health information. Each AMC’s de-identified data environment is specifically designed and operated to enable access to and analysis of de-identified data without the need for Institutional Review Board (IRB) oversight, approval, or an exemption confirmation. Given these measures, informed consent and IRB review were not required for this study.

## Data Availability

This study involves the analysis of de-identified Electronic Health Record (EHR) data via the nference nSights Federated Clinical Analytics Platform (nSights). Data shown and reported in this manuscript were extracted from this environment using an established protocol for data extraction, aimed at preserving patient privacy. The data has been de-identified pursuant to an expert determination in accordance with the HIPAA Privacy Rule. Any data beyond what is reported in the manuscript, including but not limited to the raw EHR data, cannot be shared or released due to the parameters of the expert determination to maintain the data de-identification. The corresponding author should be contacted for additional details regarding nSights.

## Code Availability

The code used in this study is not publicly available. Requests for additional information should be directed to the corresponding author.

## De-identification and HIPAA compliance certification

Prior to analysis, all EHR data were de-identified under an expert determination consistent with the Health Insurance Portability and Accountability Act (HIPAA) Privacy Rule (45 CFR §164.514(b)(1)). The de-identification methodology employed a multi-layered transformation approach to both structured and unstructured data fields.^12,13^ In structured data, direct identifiers including patient names and precise geographic locations were excluded entirely, while indirect identifiers underwent specific transformations: patient identifiers, medical record numbers, and accession numbers were replaced with one-way cryptographic hashes using confidential salts to preserve linkage across patient encounters; all dates were shifted backward by patient-specific random offsets (1–31 days) to preserve temporal relationships while obscuring exact event timing; the ZIP codes were truncated to two-digit state-level resolution; and continuous variables including age, height, weight, and body mass index were thresholded to prevent identification of extreme values (for example, ages ≥89 years transformed to ‘89+’ and BMI >40 transformed to ‘40+’). In unstructured clinical text, an ensemble de-identification system that combines attention-based deep learning models with rule-based methods achieved an estimated >99% recall for personally identifiable information (PII) detection, with detected identifiers replaced by plausible fictional surrogates.^12^

## Data Harmonization

To address heterogeneity in EHR data, we harmonized clinical variables including medications, anthropometric measurements, and diagnoses to standardized concepts. For medications, we first constructed a standardized drug concept database combining the nSights knowledge graph with RXNorm (https://www.nlm.nih.gov/research/umls/rxnorm/index.html) hierarchies to capture ingredient, brand, and dose-specific information.^14^ EHR medication records were matched using a hierarchical approach prioritizing RXNorm codes when available, followed by ingredient-level matching, and finally natural language processing and pattern matching on free-text medication orders when structured codes were absent. For anthropometric measurements (height, weight, BMI), we created a unified vocabulary from SNOMED (https://www.snomed.org/, https://athena.ohdsi.org) and LOINC (https://loinc.org/) terminologies and matched EHR measurement descriptions using standardized text matching algorithms with abbreviation expansion and synonym resolution; ambiguous mappings were resolved using OpenAI GPT-4o (https://platform.openai.com/docs/models/gpt-4o) with summary statistics as context, followed by manual verification. For diagnoses, we developed a hierarchical disease concept database from the nSights knowledge graph and matched EHR diagnosis descriptions and codes by identifying the most specific common child concept in the hierarchy. This approach enabled consistent identification of clinical entities while preserving granularity where available.

## Acknowledgements

We thank the nference engineering team for development and operation of the nSights federated AI platform.

## Author Contributions

K.M. and V.S. conceptualized and designed the study. K.M. performed weight trajectory analysis and conducted statistical analyses. G.V. developed and implemented the LLM extraction pipeline and performed downstream analysis/interpretation. A.J.V. and V.S. developed and supervised the study and provided overall conceptualization. M.C.G. provided clinical expertise and critical review of the manuscript. All authors contributed to data interpretation and approved the final manuscript.

